# Traditional Knowledge of Childhood Illnesses among the Sidama Nation, Clinical Presentation and Hematological Findings

**DOI:** 10.64898/2025.12.22.25342847

**Authors:** Serawit Deyno, Mairegu Daniel, Tsadiku Getachew, Menfese Tadesse, Hizkel Engiso, Sintayehu Fekadu, Nigatu Tuasha

**Affiliations:** School of Pharmacy, Hawassa University, Hawassa, Ethiopia; Department of Chemical Engineering, Hawassa University, Hawassa, Ethiopia; Adaare General Hospital, Hawassa, Ethiopia; Soil Resources and Watershed Management, Wondo Genet College of Forestry and Natural Resources, Hawassa University, Hawassa, Ethiopia; School of Medical Laboratory technology, Hawassa University, Hawassa, Ethiopia; Biomedical Sciences, Department of Biology, Hawassa College of Teacher Education, Hawassa, Ethiopia

**Keywords:** Case study, infant, biochemical profile, hematological profile

## Abstract

**Background:** Sidama possesses extensive indigenous knowledge regarding infant illnesses described as “*feyancho*”. Feyancho has a widespread recognition among Sidama people, though poorly documented in scientific literature. This study aimed to document the indigenous understanding of this infantile disease and examine associated clinical and biochemical profiles in the Sidama National Regional State, Ethiopia.

**Methods:** A household-based cross-sectional study was conducted in nine districts selected using systematic random sampling method. Data was collected through household surveys (n=436), focus group discussions (FGDs), and clinical assessments. Clinical and biochemical evaluations were performed on children affected by” feyancho” disease.

**Result:** The overall prevalence of “*feyancho*” was 59.52%. Most respondents (66.51%) believed that maternal health influences the occurrence of “feyancho” disease. In the study group, liver function tests showed elevated levels of SGOT (AST), ALP, and direct bilirubin in more than half of the children. Additionally, 25% presented low glucose and elevated total bilirubin. Hematological analyses revealed significant increases in LYM (+128%), EOS (+76.92%), LYM% (+79.6%), RDW-CV (+36.23%), PDW (+35.74%), and PCT (+1015.79%). Conversely, decreases were observed in NEU (–29.05%), NEU% (–36.78%), RBC (–13.26%), HGB (–28.07%), HCT (–29.36%), MCV (–20.73%), and PLT (–25.67%).

**Conclusion:** The Sidama people’s indigenous knowledge of infantile diseases remains an integral part of local health practices. Clinical and biochemical findings suggest conditions such as microcytic anemia, inflammation, or chronic infections. Safeguarding this traditional knowledge and validating it scientifically is crucial, requiring more comprehensive research.

## Introduction

Indigenous knowledge systems (IKS) play a crucial role in the understanding childhood diseases and have been used for generations to diagnose and treat illnesses [1,2]. The Sidama nation commonly defines infantile diseases as “*Feyancho*”. The “*feyancho*” can be either a milder form, locally called “*Sidancho*”, or a severe form, also locally known as “*Arusicho*”. This study is the first of its kind in attempting to document this indigenous knowledge on community understanding of diseases, providing a foundation for future research in the field. By doing so, the finding of the study will be used as a starting point for the integration of IKS on identifying diseases and aligning to modern medicine understanding.

Indigenous communities follow a holistic approach to diagnosing and managing infantile illnesses, integrating cultural beliefs, environmental observations, and generational knowledge. Unlike biomedical methods that focus on specific pathogens, traditional diagnosis relies on symptom-based assessments, spiritual interpretations, and behavioural changes. Infantile illnesses are often identified through signs such as appetite loss, restlessness, body temperature fluctuations, or digestive disturbances [3].

Studying traditional understanding of infantile diseases is crucial to have a clear picture of the merits and demerits. Understanding these practices enables healthcare providers to foster trust, bridge cultural gaps, and integrate effective remedies into formal healthcare, ultimately enhancing infant health outcomes while preserving cultural heritage [4].

There has been no scientific investigation into how these diseases are understood by the local community. To fill this gap, the present study aimed to document the IKS related to “*feyancho*” among selected rural communities in the Sidama National Regional State (SNRS) and explored how people in the study area understand this infant disease and examined clinically and biochemically.

## Materials and Methods

### The study area

The study was conducted in nine (9) Woredas (Districts) of the SNRS stratified based on the agro-climatic zones of the region. Two kebeles (the smallest administrative unit) were randomly selected from each woreda. Three different agro-climatic zones exist in Sidama [5]. These are “*Kolla*”, “*Woina Dega*” and “*Dega*” agro-climatic zones. ‘*Kolla’* is characterized by a dry, hot tropical climate area and lies between 500 and 1500 meters above sea level. It receives an annual rainfall of 400-800 mm, with a mean annual temperature of 20-25 °C and constitutes 30% of the total area. The ‘*Woina Dega*’ is characterized by a moist to humid, warm subtropical climate and is found between 1500 – 2500 meters above sea level, with an annual rainfall of 1000 – 1800 mm. This agro-climatic zone has a mean annual temperature of 15-20 °C and constitutes 54% of the total area of the region. The ‘Dega’ agro-climatic zone is known with its wet, cool temperate climate.” *Dega*” has an elevation of 2500 to 3500 meters above sea level and receives an annual rainfall of 1200 to 1800 mm, and the mean annual temperature is 10-15 °C and constitutes only 16% of the total area of the region [6].

The nine woredas included in the present study were Borricha, Daraara and Hawassa Zuria woredas representing *Qolla’* (dry, hot tropical climate); Haweela, Daalle, Daarra woredas from *Woina Dega’* (moist to humid, warm subtropical climate) and Hula, Bursa and Arbagoona from *Dega’* (wet, cool temperate climate). From each of the selected districts, two kebeles (a total of 18 kebeles) were randomly selected for the study. The sampling units are households from the selected kebeles. The study area is presented in Figure 1.

**Figure 1:**
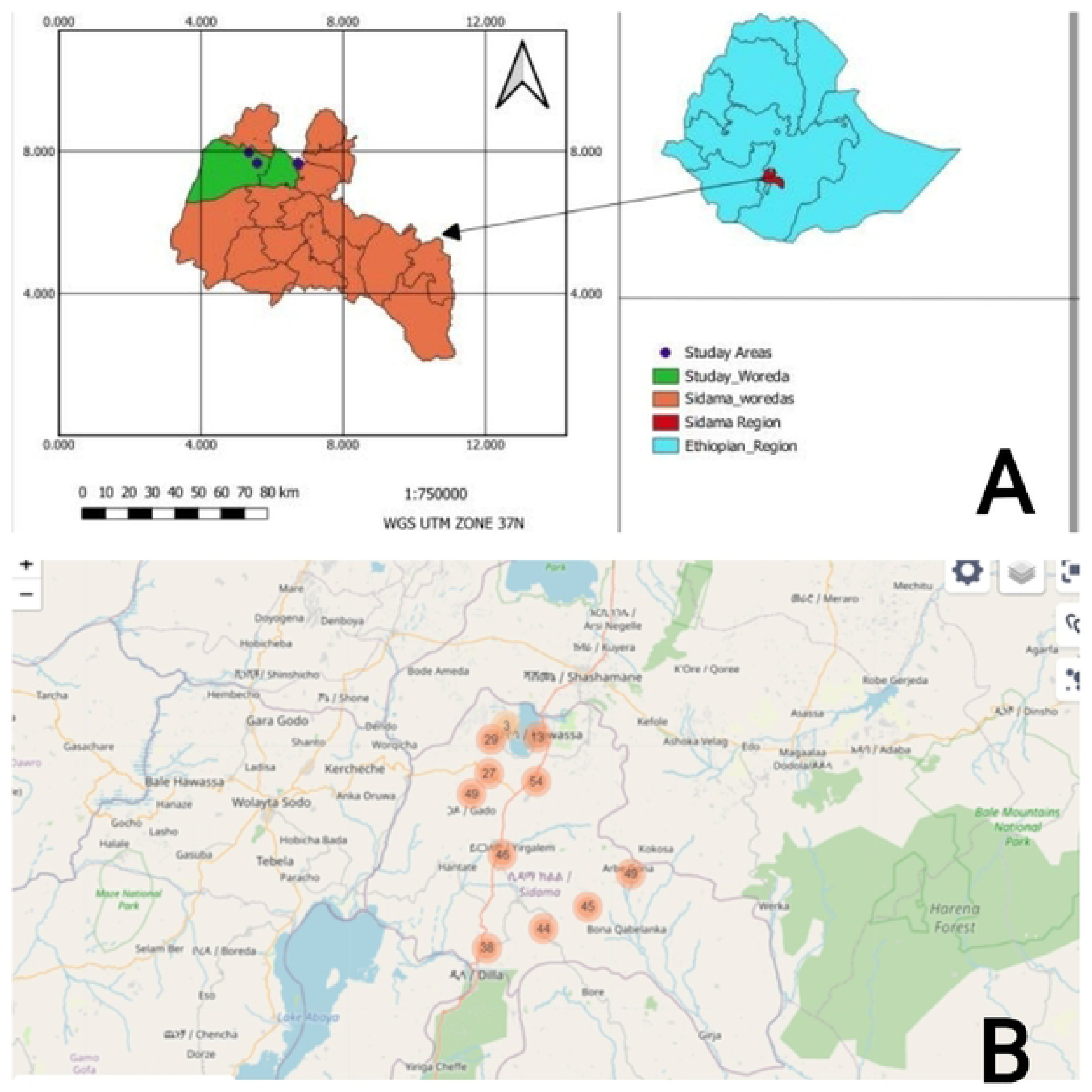
Map showing the woredas (districts) included in the study from Sidama National Regional State, Ethiopia, A drawn using QGIS and B-Kobo collect generated.

### Study population and sample collection

There is a detailed survey report based on the data collected regarding “*feyancho*” disease in SNRS, Ethiopia. The socio-demographics, understanding of diseases and symptoms of” feyancho”, disease transmission and prevention perceptions were recorded. In addition, clinical samples from children affected by the “*feyancho*” were collected for biochemical analysis. Household sample units were studied to determine the prevalence of infant diseases. The prevalence of “*feyancho*” was based on the number of total births in the sampled households.

The sample size for this study was determined based on the presence of attribute in the general population which was estimated to be at 50%. Thus, the sample size was calculated based on a single population proportion formula by taking the margin of error of 0.05 at 95% confidence level (CI), the total household unit sampled as n= (Zα/2)^2^p (1-p)/d^2^. In addition, 10% of non-response rate was considered. Accordingly, a total of 436 study participants were included in the study.

The prevalence of “feyancho”, as defined by the community, in the SNRS was determined. During this investigation, people’s perceptions and definitions of “feyancho” disease were documented. Clinical characterization and hematological parameters were evaluated from clinical samples collected from infants claimed to be affected by “feyancho”.

#### Ethics approval and consent to participate

A cross-sectional household level study was conducted from January 2023 to December 2023. Informed written assent (consent) was obtained from parents or guardians for the case study. The study was conducted in accordance with the guidelines required by the National Health Council of Ethiopia through Hawassa University Research Ethics Review Committee (Ref no: RERC 02,2022) in accordance with the Declaration of Helsinki.

#### Data Analysis

Data entry and validation were done using STATA and every possible data quality assurance approaches were followed throughout the study period. Descriptive statistics (percentages and frequency) are used to summarize the data.

## Results

### Socio-demographic characteristics

A total of 436 households were included from nine Woredas, with a diverse representation across the region. Each woreda was represented with about 9.86–12.16% participants from two kebeles. The respondents have varying educational backgrounds, ranging from no formal education to higher education levels. The respondents have diverse religious affiliations, but the majority identify as Christian Protestants. The mean age of household fathers is 45.66 (SD = 16.22) years while the median age of household mothers is 39.33 (SD = 13.50) years. The mean family size is notably larger than the mean total number of children, indicating that households also include other members beyond children, such as adults (Table 1).

**Table 1:** Socio-demographic characteristics of the study participants (N=436), 2023.

| Categorical variables |  |  |  |  |  |
| --- | --- | --- | --- | --- | --- |
| Variables | Category | N (%) | Variables | Category | N (%) |
| Education Level of Household mother | No formal education | 188 (43.12%) | Education Level of Household father | No formal education | 187 (42.89%) |
|  | Some Primary school | 184 (42.20%) |  | Some Primary school | 133 (30.50%) |
|  | Some Secondary school | 12 (2.75%) |  | Some Secondary school | 24 (5.50%) |
|  | High school complete | 22 (5.05%) |  | High school complete | 45 (10.32%) |
|  | Diploma | 19 (4.36%) |  | Diploma | 18 (4.13%) |
|  | Bachelor’s degree | 4 (0.92%) |  | Bachelor’s degree | 17 (3.90%) |
|  | Master’s degree | 1 (0.23%) |  | Master’s degree | 1 (0.23%) |
| Religion of Household father | Christian-protestant | 352 (80.73%) | Religion of Household mother | Christian-protestant | 346 (79.36%) |
|  | Apostolic | 40 (9.17%) |  | Apostolic | 44 (10.09%) |
|  | Catholics | 15 (3.44%) |  | Catholics | 14 (3.21%) |
|  | Muslim | 11 (2.52%) |  | Muslim | 11 (2.52%) |
|  | Adventist | 9 (2.06%) |  | Adventist | 9 (2.06%) |
|  | Christian-orthodox | 6 (1.38%) |  | Christian-orthodox | 7 (1.61%) |
|  | Atheist | 2 (0.46%) |  | Atheist | 1 (0.23%) |
|  | Traditional | 1 (0.23%) |  | Traditional | 1 (0.23%) |
| Continuous variables |  |  |  |  |  |
|  | Mean | Median | Mode | Standard Deviation |  |
| Family Size | 6.03 | 6.0 | 7.0 | 2.19 |  |
| No of Children | 5.21 | 4 | 2 | 3.27 |  |
| No of Male | 2.72 | 2 | 2 | 2.01 |  |
| No of female | 2.49 | 2 | 2 | 1.82 |  |

## Prevalence of the infant diseases (“feyancho”)

The total number of male and female children in the surveyed households was 1,186 and 1,084, respectively. Among the reported cases, 658 male children and 693 female children were believed to have “*feyancho”*, categorized as either *Sidancho* or *Arusicho*. The prevalence among male children was 55.48%, while among female children, it was 63.93%. The total prevalence across all children was calculated at 59.52%. These prevalence rates are derived from the total number of children surveyed, while household-level prevalence details are provided in Table 2.

**Table 2:** Prevalence of “*feyancho*” as defined by the community among study participants (N=436), 2023.

| Variable | Category | Frequency (%) |
| --- | --- | --- |
| Households have children with a mild “*feyancho*” (*Sidancho*) | Yes | 275 (67.24%) |
|  | No | 134 (32.76%) |
| Households have children with a severe “*feyancho*” (*Arusicho*) | Yes | 124 (58.22%) |
|  | No | 89 (20.41%) |
| Household belief on route of transmission of “*feyancho*” | Mother to child (MTC) | 250 (57.34%) |
|  | Breastfeeding | 176 (40.37%) |
|  | Contact | 63 (14.45%) |
|  | Genetic | 41 (9.4%) |
|  | Airborne | 2 (0.46%) |
| Age of infants affected by “*feyancho*” | Up to 1 year | 218 (50%) |
|  | Up to 5 months | 73 (16.74%) |
|  | Up to 3 months | 62 (14.22%) |

### Symptoms and characteristics of the diseases as defined by the community

The symptoms vary in frequency, with skin rashes being the most common and hair loss the least common among those believed to be affected by the milder form of “*feyancho*” (*Sidancho*). Significant portion of individuals report abdominal pain as one of major symptoms (Figure 2).

**Figure 2:**
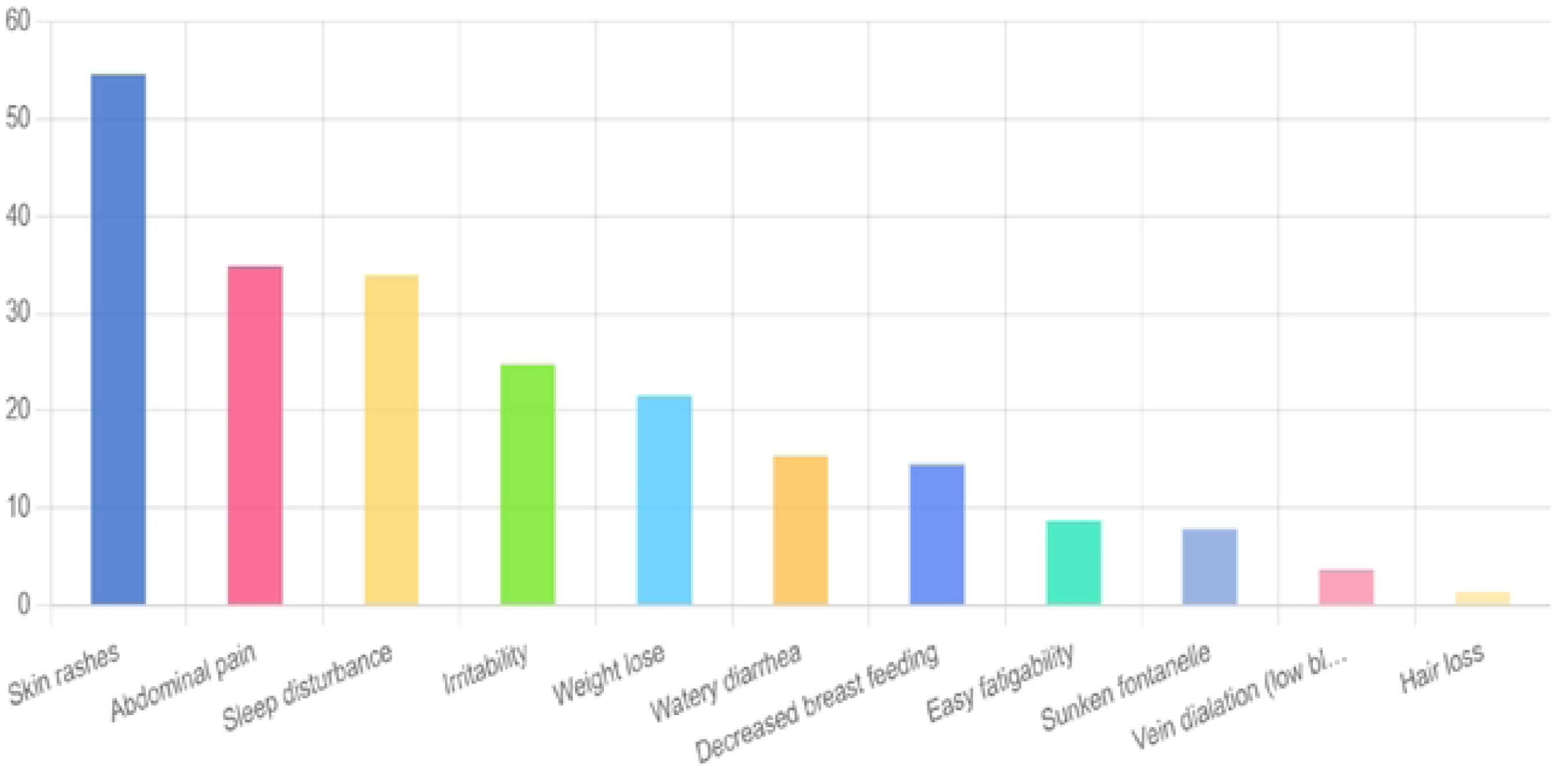
Frequency of common symptom of the mild “*feyancho*” (*Sidancho*) as described by the community.

The severe form of “*feyancho*” (*Arusicho*) is associated with several symptoms, which may include physical symptoms, psychological symptoms and cultural or spiritual implications: in some contexts, the symptoms might be interpreted as being linked to cultural or spiritual beliefs, potentially influencing the perception and treatment of the symptoms. Most reported symptom during the severe form of “*feyancho*” (*arusicho*) is that of sunken fontanelle (Figure 3).

**Figure 3:**
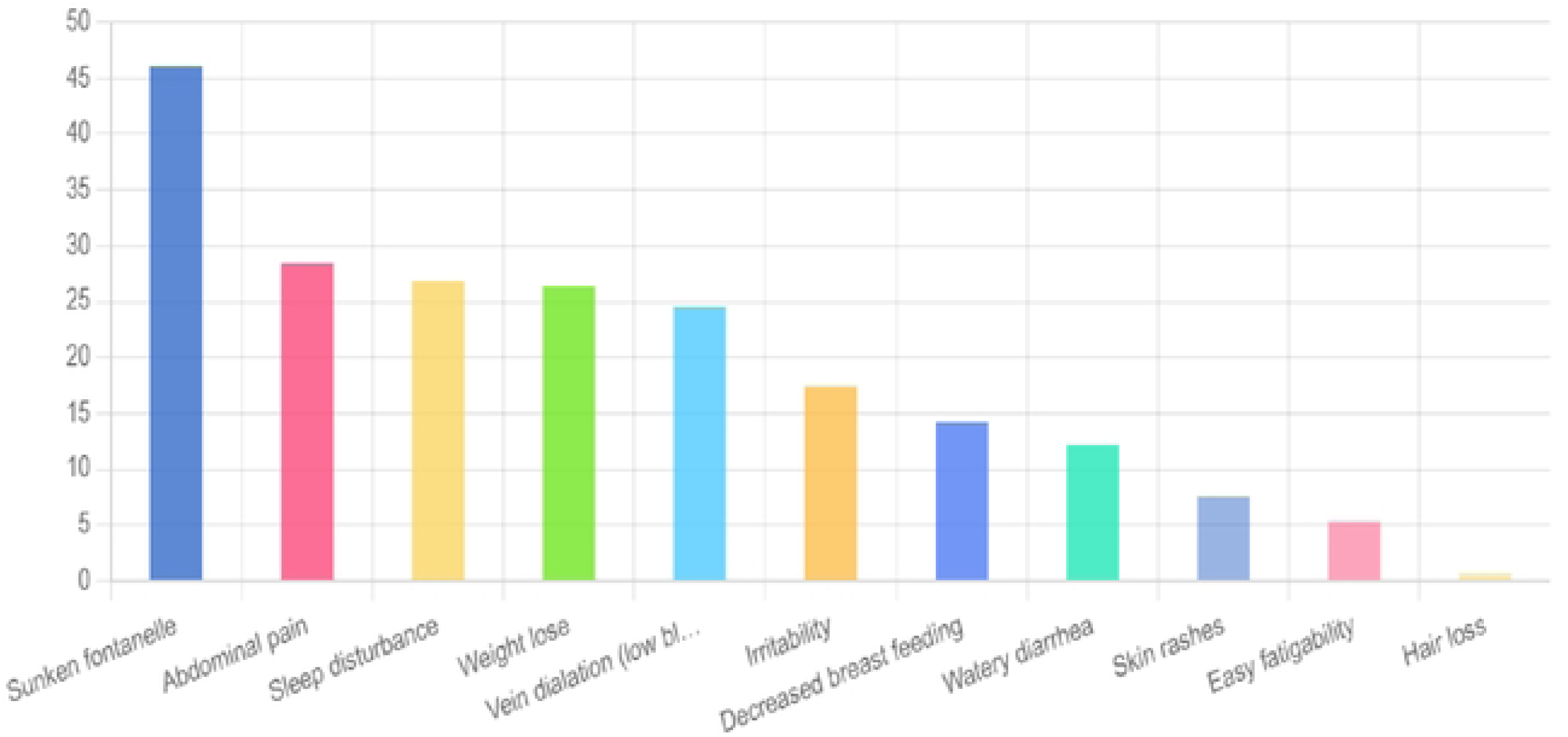
Frequency of common symptom of the severe form of “*feyancho*” (*arusicho*) as described by the community.

According to the participants, *sidancho* or *arusicho* primarily affects very young children, with the highest prevalence observed in infants up to one year old, as reported by 218 respondents (50%). Within this age range, 19 respondents (4.36%) specifically identified children up to 8–10 months, while 73 respondents (16.74%) noted cases in infants between 3–5 months. Additionally, 62 respondents (14.22%) indicated that children up to 3 months were affected (Table 2). The data highlights that early infancy is the most vulnerable period for these diseases, emphasizing the need for targeted health interventions during this stage.

### Study participants’ belief on the mechanisms of transmission of the diseases

Most participants (N=290; 66.51%) believed that a mother affected by “*feyancho*” increases the likelihood of her children being affected. In contrast, 73 respondents (16.74%) did not share this belief. These findings suggest that most participants recognize a potential hereditary or environmental link between a mother’s condition and her children’s susceptibility to “*feyancho*”.

The participants identified several possible ways through which “*feyancho*” may be transmitted. The most believed mode of transmission was direct transmission from mother to child, with 250 respondents (57.34%) supporting this view. Similarly, a significant portion of respondents (176 cases, 40.37%) believed that transmission occurs through breastfeeding, indicating concerns about the role of maternal feeding practices in the spread of the disease. Overall, the findings indicate that most participants believe maternal transmission, particularly through direct contact and breastfeeding, is the primary way “*feyancho*” spreads among children (Table 2).

### Community’s perception about the symptoms and amelioration of “*feyancho*”

Focused group discussions (FDGs) were conducted with community participants to complement the data obtained from structured interviews. A total of eight interview sessions were carried out to explore people’s understanding of the disease condition known as *”feyancho”*. The summaries of these interviews provide insights into participants’ perceptions and statements and are complementary with the interview findings.

Sidama nation categorizes “*feyancho*” into two types: *arusicho* (a severe form) and *sidancho* (a milder form). “*Feyancho”* is characterized by a range of symptoms and conditions. Severe cases are characterized by skin peeling, sunken fontanels, emaciation, and stomach issues. Milder conditions involve symptoms such as stomach-aches, diarrhoea, restlessness, and minor skin rashes. The amelioration of the conditions is characterized by purging of green diarrhoea, disappearance of rushes, good sleeping, and fontanels normalized, resumes normal breastfeeding, physical betterment and fattening, the stiffened belly begins to soften and so on.

### Clinical characteristics of children claimed to have “*feyancho”*

Examinations were conducted by physicians to clinically define the “*feyancho” (arusicho/sidancho)*. It included data on the place of birth, previous episodes, symptom resolution, and gender distribution. The table also covered medical evaluation and hospitalization history, maternal employment, respiratory and environmental exposures, and immunization status. Various symptoms and conditions were detailed, such as fever, sweating, chills, cough, wheezing, shortness of breath, chest pain, runny nose, throat issues, difficulty swallowing, ear pain, red draining eye, muscle ache, joint pain, swollen glands, rash, stiffened neck, seizures, vomiting, diarrhea, bloody urine, yellow skin/eyes, weight loss, irritability, skin rashes, and hair loss (Table 3).

**Table 3:** Clinical characteristics of *“feyancho” (sidancho/ arusicho)* in SNRS, 2023.

| Characteristics | Options | Frequency (%) | Characteristics | Options | Frequency (%) |
| --- | --- | --- | --- | --- | --- |
| Place of birth | Facility-based | 11(84.62) | Muscle ache | Yes | 0 |
|  |  |  |  | No | 8(61.54) |
|  | Home delivery | 2(15.38) |  | UK | 5(38.46) |
| Ever have similar episode | Yes | 3(23.08) | Joint pain/swelling | Yes | 0 |
|  | No | 10(76.92) |  | No | 13(100) |
| Similar case in the vicinity | Yes | 3(23.08) | Enlarged/swollen glands | Yes | 1(7.69) |
|  | No | 10(76.92) |  | No | 12(92.31) |
|  | Yes | 3(23.08) | Rash | Yes | 9(69.23) |
| Symptom-resolved without intervention | No | 10(76.92) |  | No | 4(30.77) |
| Sex | Male | 9(69.23) | Red draining eye | Yes | 1(7.69) |
|  | Female | 4(30.77) |  | No | 12(92.31) |
| Ever evaluated by physician before | Yes | 1(7.69) | Bloody urea | Yes | 0 |
|  | No | 12(92.31) |  | No | 13(100) |
| Ever hospitalized | Yes | 1(7.69) | Yellow skin/eye | Yes | 0 |
|  | No | 12(92.31) |  | No | 13(100) |
| Mother works outside home | Yes | 3(23.08) | T<sup>0</sup>c raised | Yes | 0 |
|  | No | 10(76.92) |  | No | 3(23.08) |
| Ever have respiratory distress | Yes | 0 | Wheezing bronchi | UK | 10(76.92) |
|  | No | 13(100) |  | Yes | 0 |
| Ever exposed to soil | Yes | 4(30.77) | Mucosal lesions | No | 2(15.38) |
|  | No | 9(69.23) |  | UK | 11(84.62) |
| Have pets in the same house | Yes | 5(38.46) | Altered mental status | Yes | 2(15.38) |
|  | No | 8(61.54) |  | No | 11(84.62) |
| Ever have contacts with pets | Yes | 2(15.38) | Neurological abnormalities | Yes | 0 |
|  | No | 11(84.62) |  | No | 13(100) |
| Ever have contacts with rodents | Yes | 4(30.77) | Hepatosplenomegaly | Yes | 0 |
|  | No | 9(69.23) |  | No | 13(100) |
| Immunized | Yes | 13(100) | Jaundice | Yes | 0 |
|  | No | 0 |  | No | 4(30.77) |
| Ever used medication including OTC | Yes | 7(53.85) | Excessive crying | UK | 9(69.23) |
|  | No | 6(46.15) |  | Yes | 0 |
| Ever have depressed fontanelle | Yes | 0 | Constipation | No | 13(100) |
|  | No | 6(46.15) |  | Yes | 9(69.23) |
|  | Unknown | 7(53.85) |  | No | 4(30.77) |
| Current fever | Yes | 7(53.85) | Failure to thrive | Yes | 1(7.69) |
|  | No | 6(46.15) |  | No | 12(92.31) |
| Current sweet | Yes | 10(76.92) | Reduced breast Feeding | Yes | 5(38.46) |
|  | No | 3(23.08) |  | No | 8(61.54) |
| Current chills | Yes | 2(15.38) | Abdominal pain | Yes | 4(30.77) |
|  | No | 11(84.62) |  | No | 9(69.23) |
| Current cough | Yes | 7(53.85) | Watery diarrhea | Yes | 0 |
|  | No | 6(46.15) |  | No | 5(38.46) |
| Current cough with sputum | Yes | 0 | Weight loss | UK | 8(61.54) |
|  | No | 13(100) |  | Yes | 10(76.92) |
| Wheezing | Yes | 0 | Irritability | No | 3(23.08) |
|  | No | 12(92.31) |  | Yes | 11(84.62) |
|  | Unknown | 1(7.69) |  | No | 2(15.38) |
| Shortness of breath | Yes | 0 | Skin rashes | Yes | 10(76.92) |
|  | No | 12(92.31) |  | No | 3(23.08) |
| Chest pain | Yes |  | Hair loss | Yes | 10(76.92) |
|  | No | 7(53.85) |  | No | 3(23.08) |
|  | Unknown | 6(46.15) |  | Yes | 2(15.38) |
| Runny nose | Yes | 0 | Stiff neck | No | 11(84.62) |
|  | No | 13(100) |  | Yes | 1(7.69) |
| Nose throat | Yes | 2(15.38) | Seizures | No | 12(92.31) |
|  | No | 11(84.62) |  | Yes | 2(15.38) |
| Difficulty swallowing | Yes | 1(7.69) | Vomiting | No | 11(84.62) |
|  | No | 12(92.31) |  | Yes | 7(53.85) |
| Ear pain | Yes | 1(7.69) |  | No | 6(46.15) |
|  | No | 8(61.54) | Vomiting with blood | Yes | 0 |
|  | Unknown | 4(30.77) |  | No | 13(100) |

### Clinical manifestations of the “*feyancho*” disease

A total of 12 clinical cases were investigated. The study found that creatinine and serum glutamic-pyruvic transaminase (SGPT) / alanine aminotransferase (ALT) levels were within normal ranges for all participants. However, blood urea levels were elevated in one participant (8.33%). A significant percentage of participants exhibited elevated levels of Serum Glutamic-Oxaloacetic Transaminase (SGOT) aspartate aminotransferase (AST) (58.33%), Alkaline Phosphatase (ALP) (91.66%), and Direct Bilirubin (DB) (58.33%).

A notable concern is the presence of hypoglycemia in 25% of the participants. While cholesterol levels were within a healthy range across the participants, bilirubin findings (both total and direct) pointed to mild jaundice in some cases. Overall, this study highlights that while most participants exhibit healthy biochemical profiles, a subset presented with abnormal liver enzyme levels, hypoglycemia, and elevated bilirubin (Table 4).

**Table 4:** Clinical chemistry of samples collected from children with Feyancho disease, 2023.

| Variable | Range in study | Mean | Range less than 1year | %below normal | %above normal | Laboratory range |
| --- | --- | --- | --- | --- | --- | --- |
| Creatinine (mg/dl) | 0.21-0.39 | 0.31 | 0.2-0.4 | 0 | 0 | 0.9-1.3 |
| Urea (mg/dl) | 6.3-18.2 | 12.21 | 5 to 18 | 0 | 8.33 | 16.6-48.5 |
| SGPT U/L | 11.3-50.9 | 24.72 | 7-56 | 0 | 0 | 0-41 |
| SGOT U/L | 5-40 | 52.69 | 5-40 | 0 | 58.33 | 0-40 |
| ALP U/L | 244.8 -<br>1237.9 | 701.9 | 81.9 -<br>350.3 | 0 | 91.66 | 120-460 |
| TC (mg/dl) | 70.9 -147.9 | 121.92±29.<br>193 | <170 | 0 | 0 | 0-200 |
| Glucose (mg/dl) | 3.4 -108.8 | 64.17±<br>31.18 | 60-200 | 25 | 0 | 74-109 |
| Albumin (g/l) | 3.52-4.66 | 4.35±38.84 | 3.4-5.2 | 0 |  | 3.5-5.2 |
| TB (mg/dl) | 0.2- 3.28 | 1.15± 1.01 | <2.0 | 0 | 25 | 0.2-1.0 |
| DB (mg/dl) | 0.23 -4.15 | 1.40±1.36 | <0.3 | 0 | 58.33 | 0-0.2 |
| TP (mg/dl) | 6.6 -8.6 | 7.83± 0 .58 | 4.5 -7.0 |  |  | 6.6-8.7 |

### Hematological parameters of the “*feyancho*” disease

The findings from the hematological study may indicate the children with underlying anemia and some immune activity, potentially linked to infections or inflammatory conditions. The study findings indicate that white blood cell (WBC) and neutrophil counts among the participants were within the normal range. However, the mean lymphocyte percentage was notably higher than normal. Monocyte and eosinophil counts were largely within normal limits, with eosinophil levels slightly elevated at the upper end of the normal range. Basophil levels were on the lower end of normal. The lower-than-normal neutrophil percentage likely correlates with the relatively higher lymphocyte count, reinforcing the possibility of underlying viral infections (Table 5).

**Table 5:** Investigation of the hematological parameters of the samples collected from children with Feyancho disease, 2023.

| *Variable* | *Min*<br>*value* | *Max*<br>*value* | *Observed*<br>*Mean* | *Normal*<br>*Mean* | *Difference* | *%*<br>*Difference* | *Deviation* |
| --- | --- | --- | --- | --- | --- | --- | --- |
| *WBC*<br>($\times 10^3/\mu\text{L}$) | 4.560 | 18.21 | 8.84 | 7.5 | +1.34 | +17.87% | Elevated |
| *NEU*<br>($\times 10^3/\mu\text{L}$) | 1.300 | 6.86 | 3.37 | 4.75 | -1.38 | -29.05% | Decreased |
| *LYM*<br>($\times 10^3/\mu\text{L}$) | 2.760 | 7.33 | 4.56 | 2.0 | +2.56 | +128.00% | Elevated |
| *MON*<br>($\times 10^3/\mu\text{L}$) | 0.200 | 0.93 | 0.42 | 0.6 | -0.18 | -30.00% | Decreased |
| *EOS*<br>($\times 10^3/\mu\text{L}$) | 0.050 | 3.24 | 0.46 | 0.26 | +0.20 | +76.92% | Elevated |
| *BAS*<br>($\times 10^3/\mu\text{L}$) | 0.020 | 0.02 | 0.02 | 0.1 | -0.08 | -80.00% | Decreased |
| *NEU %* | 21.400 | 50.80 | 37.93 | 60.0 | -22.07 | -36.78% | Decreased |
| *LYM %* | 36.000 | 70.60 | 53.88 | 30.0 | +23.88 | +79.60% | Elevated |
| *MON %* | 2.400 | 7.00 | 4.78 | 5.0 | -0.22 | -4.40% | Normal |
| *EOS %* | 0.700 | 17.80 | 3.52 | 3.5 | +0.02 | +0.57% | Normal |
| *BAS %* | 0.100 | 0.50 | 0.29 | 0.5 | -0.21 | -42.00% | Decreased |
| *RBC*<br>($\times 10^6/\mu\text{L}$) | 2.510 | 4.98 | 4.12 | 4.75 | -0.63 | -13.26% | Decreased |
| *HGB*<br>(g/dL) | 4.600 | 14.00 | 10.07 | 14.0 | -3.93 | -28.07% | Decreased |
| *HCT (%)* | 15.400 | 40.60 | 29.67 | 42.0 | -12.33 | -29.36% | Decreased |
| *MCV (fL)* | 54.500 | 81.80 | 71.34 | 90.0 | -18.66 | -20.73% | Decreased |
| *MCH (pg)* | 16.200 | 28.20 | 24.24 | 29.5 | -5.26 | -17.83% | Decreased |
| *MCHC*<br>(g/dL) | 29.800 | 37.20 | 33.86 | 34.0 | -0.14 | -0.41% | Normal |
| *RDW-CV*<br>(%) | 13.300 | 25.90 | 17.71 | 13.0 | +4.71 | +36.23% | Elevated |
| *RDW-SD*<br>(fL) | 39.700 | 55.70 | 47.12 | 42.5 | +4.62 | +10.87% | Elevated |
| *PLT*<br>($\times 10^3/\mu\text{L}$) | 17.000 | 501.00 | 223.0 | 300 | -77.0 | -25.67% | Decreased |
| *MPV (fL)* | 7.100 | 12.40 | 9.34 | 9.5 | -0.16 | -1.68% | Normal |
| *PDW (fL)* | 14.300 | 16.40 | 15.61 | 11.5 | +4.11 | +35.74% | Elevated |
| *PCT (%)* | 0.120 | 4.55 | 2.12 | 0.19 | +1.93 | +1015.79% | Elevated |
| *PLCR* | 0.172 | 0.44 | 0.25 | 0.28 | -0.03 | -10.71% | Normal |

## Discussion

Illnesses in newborns typically range from mild skin rashes and common colds to more severe infections and genetic defects. While some are easily managed at home, others may require prompt medical attention. “*Feyancho*” among Sidama nation is well characterized symptomatically and almost exclusively managed at home. Yet, its western equivalence has not yet been established. To establish the etiological manifestations, communities’ description of the diseases, signs and symptoms, clinical samples were analyzed. This study, the first in its kind, has found out that the prevalence of “*feyancho*” disease is higher among females (63.93%) than males (55.48%). Clinical evaluations of “*feyancho*” cases revealed the possibility of microcytic anemia, high lymphocytes, low neutrophils, increased RDW (Red Cell Distribution Width), anisocytosis, and abnormal platelet indices suggest iron deficiency, inflammation, or chronic infection.

Slightly higher prevalence among female might indicate that females are either more susceptible to “*feyancho*” disease possibly due to biological or environmental factors. A previous study conducted in the region found that all children tested were positive for fungal elements (dermatophytosis) in microscopy with 97.6% exhibited growth in culture [7]. The skin manifestations observed in the current study participants may agree with the report.

Regarding disease classification, participants categorized “*feyancho*” into two types: *sidancho* (a milder form) and *arusicho* (a severe form). *Arusicho* is associated with more severe clinical manifestations mainly manifested as sunken fontanelles and ultimately leads to death if left untreated with traditional treatments.

A sunken fontanel can be caused by dehydration, kwashiorkor (severe protein deficiency), failure to thrive (poor growth in children), and serious medical conditions like toxic megacolon and diabetes insipidus. The study of Tswana-speaking urbanites’ cultural perceptions of diarrhoeal disease in infants is often linked to sorcery, with concepts like “phogwane” (sunken fontanelle) and “kokwana” (intestinal snake) used to explain its origins [8]. This concept of a sunken fontanelle overrides among several communities and is attributed to various understandings and treatment aspects, and culturally informed and plausible approaches are important to such conditions [9].

A significant majority (66.51%) of respondents believed that mothers affected by “*feyancho*” increased the likelihood of their children contracting the disease. Maternal-to-child transmission was considered the primary route (57.34%), followed by transmission through breastfeeding (40.37%) and direct contact among children (14.45%). Genetic and airborne transmissions were believed to play a minimal role. This indicates that cultural beliefs about transmission heavily influence local health perceptions and may impact preventive health behaviors.

Medical evaluations provided additional insights into the disease’s presentation. Notably, symptoms such as excessive crying, weight loss, and irritability were commonly observed in affected children. Despite the widespread community belief in maternal transmission, clinical examinations did not find strong correlations with maternal health conditions. Furthermore, hospitalization rates among affected children were low, suggesting either underreporting of severe cases or limited access to healthcare facilities.

Laboratory analysis revealed that a significant portion of participants exhibited elevated SGOT, ALP, and direct bilirubin (DB) levels, suggesting potential liver dysfunction or stress, possibly linked to the underlying conditions affecting liver function. In contrast, creatinine and urea levels remained mostly within normal ranges, indicating no major kidney issues. However, the slight elevation in urea observed in some participants warrants further assessment of hydration status or early signs of kidney function impairment. Presence of hypoglycemia in infants can negatively impact neurological development if left unaddressed. While cholesterol levels were within a healthy range across participants, bilirubin findings (both total and direct) pointed to mild jaundice in some cases—an expected condition in infants but one that requires close monitoring. Overall, this study highlights that while most participants exhibit healthy biochemical profiles, a subset presents with abnormal liver enzyme levels, hypoglycemia, and elevated bilirubin, warranting further investigation to rule out any underlying health concerns.

The haematological analysis revealed several key findings. Red blood cell (RBC) parameters suggest a prevalence of anaemia among participants, as evidenced by low RBC count, haemoglobin, and haematocrit levels. The reduced mean corpuscular volume (MCV) and mean corpuscular haemoglobin (MCH) indicate microcytic anaemia, commonly associated with iron deficiency [10]. Despite these findings, the mean corpuscular haemoglobin concentration (MCHC) remained within normal limits, suggesting that while red blood cells were smaller; their haemoglobin concentration was not significantly altered. An elevated red cell distribution width (RDW) further supports the presence of anisocytosis, a condition characterized by varying RBC sizes, often seen in nutritional deficiencies [11].

Platelet counts were within the normal range, indicating no major clotting disorders, though platelet distribution width (PDW) and plateletcrit (PCT) were slightly elevated, suggesting variations in platelet size and potential changes in platelet production. Additionally, elevated lymphocyte levels may point to underlying viral infections or chronic inflammatory conditions [12]. Despite slight increases in PDW and PCT, platelet function appears normal, as overall platelet counts remain within the expected range. Lastly, eosinophil levels show mild elevations in some cases, which could indicate allergic reactions or parasitic infections among certain participants [13].

## Conclusion

This study provided valuable insights into the characteristics and health conditions associated with “*feyancho*”. The findings underscore the need for targeted interventions to address both healthcare access and community health education to mitigate the impact of “*feyancho*”. Given the high prevalence of “*feyancho*” and the strong cultural beliefs surrounding its transmission, community-based health education programs should be implemented. These programs should focus on improving awareness of evidence-based disease transmission and preventive measures. Currently, cases are managed through traditional methods. Strengthening collaboration between healthcare providers and local communities will be very crucial in improving child health outcomes in the region. Further epidemiological studies that integrate medical diagnostics with community perceptions could help bridge the knowledge gap and inform public health interventions.

## List of abbreviations

ALP: Alkaline Phosphatase
ALT: Alanine Aminotransferase
AST: Aspartate Aminotransferase
CAM: Complementary and Alternative Medicine
DB: Direct Bilirubin
EOS: Eosinophils
FGD: Focus Group Discussion
GPS: Global Positioning System
HCT: Hematocrit
HGB: Hemoglobin
IKS: Indigenous Knowledge Systems
LYM: Lymphocytes
MCH: Mean Corpuscular Hemoglobin
MCHC: Mean Corpuscular Hemoglobin Concentration
MCV: Mean Corpuscular Volume
MPV: Mean Platelet Volume
NEU: Neutrophils
OTC: Over the Counter
PCT: Platelets
PDW: Platelet Distribution Width
PhD: Doctor of Philosophy
PIM: Pediatric Integrative Medicine
PLCR: Platelet Large Cell Ratio
PLT: Platelets
RDW-CV: Red Cell Distribution Width-Coefficient of Variation
RDW-SD: Red Cell Distribution Width-Standard Deviation
SGOT: Serum Glutamic-Oxaloacetic Transaminase
SNRS: Sidama National Regional State
SPGT: Serum Glutamic-Pyruvic Transaminase
TB: Total Bilirubin
TC: Total Cholesterol
TP: Total Protein
WBC: White Blood Cells

## Clinical trial number

Not applicable

## Acknowledgements

The authors would like to extend their gratitude to the Research and Coordination Vice President of the Hawassa University for funding this project.

## Author contributions

SD: Conceptualization, Data Curation, Formal Analysis, Funding Acquisition, Investigation, Methodology, Project Administration, Resources, Software, Supervision, Validation, Visualization, Writing – Original Draft Preparation, and Writing – Review & Editing

MD: Conceptualization, Resources, Software, Supervision, Validation, Visualization, and Writing – Review & Editing

TG: Conceptualization, Data Curation, Formal Analysis, Funding Acquisition, Investigation, Methodology, Project Administration, Resources, Software, Supervision, Validation, Visualization, Writing – Original Draft Preparation, and Writing – Review & Editing

MT: Resources, Supervision, Validation, Visualization, and Writing – Review & Editing

HE: Resources, Supervision, Validation, Visualization, and Writing – Review & Editing

SF: Conceptualization, Funding Acquisition, Investigation, Methodology, Project Administration, Resources, Supervision, Validation, Visualization, and Writing – Review & Editing

NT: Conceptualization, Data Curation, Funding Acquisition, Investigation, Project Administration, Resources, Supervision, Validation, Visualization, Writing – Original Draft Preparation, and Writing – Review & Editing

## Data availability

Data are provided within the manuscript, or additional data will be available from the corresponding author upon request.

## Ethics declarations

### Ethics approval and consent to participate

The study was conducted in accordance with the guidelines required by the National Health Council of Ethiopia through Hawassa University Research Ethics Review Committee (Ref no: RERC 02,2022) in accordance with the Declaration of Helsinki. The participants provided their written informed consent to participate in this study.

### Consent for publication

Not applicable.

## References

1. Towns AM, Mengue Eyi S, Van Andel T. Traditional medicine and childcare in Western Africa: Mothers’ knowledge, folk illnesses, and patterns of healthcare-seeking behavior. PLoS One. 2014;9. doi:10.1371/journal.pone.0105972

2. Hailu F, Cherie A, Gebreyohannis T, Hailu R. Determinants of traditional medicine utilization for children: A parental level study in tole district, Oromia, Ethiopia. BMC Complement Med Ther. 2020;20. doi:10.1186/s12906-020-02928-1

3. Burgard P. A holistic approach to the patients/Families with inborn errors of metabolism. J Mother Child. 2020;24: 65–72. doi:10.34763/jmotherandchild.20202402si.2004.000010

4. James PB, Gyasi RM, Kasilo OMJ, Wardle J, Bah AJ, Yendewa GA, et al. The use of traditional medicine practitioner services for childhood illnesses among childbearing women: a multilevel analysis of demographic and health surveys in 32 sub-Saharan African countries. BMC Complement Med Ther. 2023;23. doi:10.1186/s12906-023-03972-3

5. Abebe T, Sterck FJ, Wiersum KF, Bongers F. Diversity, composition and density of trees and shrubs in agroforestry homegardens in Southern Ethiopia. Agroforestry Systems. 2013;87. doi:10.1007/s10457-013-9637-6

6. Elbakidze M, Gebrehiwot M, Angelstam P, Yamelynets T, Surová D. Defining priority land covers that secure the livelihoods of urban and rural people in Ethiopia: A case study based on citizens’ preferences. Sustainability (Switzerland). 2018;10. doi:10.3390/su10061701

7. Haro M, Alemayehu T, Mikiru A. Dermatophytosis and its risk factors among children visiting dermatology clinic in Hawassa Sidama, Ethiopia. Sci Rep. 2023;13. doi:10.1038/s41598-023-35837-7

8. Booyens JH. [Aspects of the popular attitude about diarrhea among Tswana-speaking urbanites]. Curationis. 1989;12.

9. Pachter LM, Weller SC, Baer RD, de Alba Garcia JEG, Glazer M, Trotter R, et al. Culture and Dehydration: A Comparative Study of Caída de la Mollera (Fallen Fontanel) in Three Latino Populations. J Immigr Minor Health. 2016;18. doi:10.1007/s10903-015-0259-0

10. Mohammed S, Mousa A., Hashim A. Role of Hypochromia and Microcytosis in theprediction of iron deficiency anemia. Minia Journal of Medical Research. 2020;31. doi:10.21608/mjmr.2022.220289

11. Salvagno GL, Sanchis-Gomar F, Picanza A, Lippi G. Red blood cell distribution width: A simple parameter with multiple clinical applications. Critical Reviews in Clinical Laboratory Sciences. 2015. doi:10.3109/10408363.2014.992064

12. Kumar V, Sharma A. Neutrophils: Cinderella of innate immune system. International Immunopharmacology. 2010. doi:10.1016/j.intimp.2010.08.012

13. O’Sullivan JA, Bochner BS. Eosinophils and eosinophil-associated diseases: An update. Journal of Allergy and Clinical Immunology. 2018;141. doi:10.1016/j.jaci.2017.09.022

